# Identification of susceptibility loci for adverse events following COVID-19 vaccination in the Japanese population: A web-based genome-wide association study

**DOI:** 10.1101/2021.11.30.21267043

**Authors:** Shun Nogawa, Hajime Kanamori, Koichi Tokuda, Kaoru Kawafune, Miyuki Chijiiwa, Kenji Saito, Shoko Takahashi

**Affiliations:** Genequest Inc., Siba 5-29-11, Minato-ku, Tokyo 108-0014, Japan; Department of Infectious Diseases, Internal Medicine, Tohoku University Graduate School of Medicine, 2-1 Seiryo-machi, Aoba-ku, Sendai 980-8574, Japan; Division of Infection Control, Tohoku University Hospital, 1-1 Seiryo-machi, Aoba-ku, Sendai 980-8574, Japan

## Abstract

The novel coronavirus disease 2019 (COVID-19) pandemic has spread rapidly worldwide. To prevent the spread of COVID-19, mRNA-based vaccines made by Pfizer/BioNTech (BNT162b1) and Moderna (mRNA-1273) have been widely used worldwide, including in Japan. Various adverse events after COVID-19 mRNA vaccinations have been reported, with differences observed among individuals. However, the analysis on the genetic background for susceptibility to side effects has been limited. In the present work, we performed genome-wide association studies (GWAS) for self-reported adverse events of COVID-19 mRNA vaccination in 4,545 Japanese individuals and identified 14 associated loci. Among these, 6p21 was associated with 37.5°C or higher fever, 38 °C or higher fever, and muscle pain. Our results may enable one to prepare for and manage side effects by knowing their susceptibility to the occurrence of adverse events. Furthermore, we obtained valuable data that can lead to the understanding of the mechanism of action of COVID-19 mRNA vaccines.

## Introduction

The novel coronavirus disease 2019 (COVID-19), caused by the severe acute respiratory syndrome coronavirus 2 (SARS-CoV-2), has spread rapidly since its emergence in December 2019 and has affected hundreds of millions of people worldwide^1^. The development and spread of safe and efficacious vaccines are now expected to be key for controlling the COVID-19 pandemic. According to the World Health Organization, several vaccines have been developed worldwide to prevent the spread of COVID-19^2^. The mRNA-based vaccines made by Pfizer/BioNTech (BNT162b1) and Moderna (mRNA-1273) are among the most widely used in Japan as well as in other parts of the world. As of November 29, 2021, the Japanese government estimates that 97.2 million people have been fully vaccinated (received two doses of either vaccine) in Japan, representing 76.7% of the country’s population^3^.

The most common side effects reported after COVID-19 mRNA vaccination are injection site reactions, fatigue, headache, and myalgia. These side effects are reported to be mild to moderate and last for a couple of days^4^. Other local and systemic side effects that occur following vaccination include swelling, chills, joint pain, fever, redness, itching, nausea, diarrhea, abdominal pain, rash outside the injection site, and vomiting^4^. Serious side effects, such as mild allergic reactions and anaphylaxis, are rare but have been reported^5,6^. Side effects are a sign of a common immune response to the vaccine and are reported to vary between age, sex, and ethnicity^7,8^. Interestingly, an elevated risk for myocarditis following COVID-19 mRNA vaccines was observed in males aged 12–29 years^9^. *HLA* alleles were also reported to be associated with adverse events following COVID-19 mRNA vaccination^10,11^. However, so far, the mechanism of the development of side effects is not clear, and there is still a lack of reports from other ethnicities.

One crucial challenge is to elucidate the factors that induce adverse events in response to the COVID-19 vaccine to understand the detailed mechanism of action. Here, we hypothesize that the individual differences in responses to the COVID-19 vaccine may be explained, in part, by genetic differences. Therefore, to clarify the possible mechanisms by which host genetic variation might affect the COVID-19 vaccine treatment response, a genome-wide association study (GWAS) was performed in the Japanese population.

## Materials and methods

### Study subjects

The data were obtained through Japanese direct-to-consumer (DTC) genetic testing services “Genequest ALL” and “Euglena MyHealth”, which are provided by Genequest Inc. (Tokyo, Japan) and Euglena Co., Ltd. (Tokyo, Japan), respectively. We asked subjects who were aged ≥ 18 years and who gave consent to participate in the study to answer internet-based questionnaires about COVID-19 vaccine adverse events. All participants provided written informed consent for the general use of their genetic data for research purposes. Prior to participating in this study, information on the study’s aim was sent to the participants and an additional study-specific agreement was obtained by opt-in. This study was conducted in accordance with the principles of the Declaration of Helsinki. The study protocol was approved by the Ethics Committee of Genequest Inc. (IRB no. 2021-0633-4) and Tohoku University Graduate School of Medicine (IRB no. 2021-1-469).

### DNA sampling, genotyping, quality control, and genotype imputation

Saliva samples were collected, stabilized, and transported using an Oragene DNA Collection Kit (DNA Genotek Inc., Ottawa, Ontario, Canada) or GeneFix Saliva DNA Collection (Cell Projects Ltd, Harrietsham, Kent, UK). Genotype analysis was performed using: Illumina Infinium Global Screening Array v1+ Custom BeadChip (Illumina, San Diego, CA, USA), which contains 704,589 markers; Infinium Global Screening Array-24 v3.0+ Custom BeadChip, which contains 655,471 markers; HumanCore-12+ Custom BeadChip, which contains 302,073 markers; HumanCore-24+ Custom BeadChip, which contains 309,725 markers; or InfiniumCore-24+ Custom BeadChip, which contains 308,500 markers. Because the analyzed single nucleotide polymorphism (SNP) sets were very different among the genotyping chips used, the subjects were divided into two groups depending on the type of genotyping chips: those analyzed by the two former chips (595,105 common markers) and those analyzed by the three later chips (289,930 common markers). These were referred to as populations A and B, respectively. We then applied the quality control and association analysis procedures separately for each cohort.

For quality control analysis, we filtered out SNP markers. The parameters were as follows: call rate per SNP < 0.95; Hardy–Weinberg equilibrium exact test *p*-value < 1 × 10^−6^; minor allele frequency < 0.01; SNPs not in autosomes. We also excluded subjects according to the following parameters: inconsistent sex information between the genotype and questionnaire; call rate per subject < 0.95; close relationship pairs determined using the identity-by-descent method (PI_HAT > 0.1875); and estimated non-Japanese ancestry. Quality control analyses were carried out using PLINK^12,13^ (version 1.90b3.42) and Eigensoft^14^ (version 6.1.3).

Genome-wide genotype imputation was performed using a pre-phasing/imputation stepwise approach implemented in EAGLE2^15^ (version 2.4) / Minimac3^16^ (version 2.0.1). The imputation reference panel was 1000 Genome Phase 3^17^ (version 5). We excluded variants with low imputation quality (R^2^ < 0.3) and low minor allele frequency (<0.05) from further analysis. Finally, we used the dosage data for the common 5,930,410 variants for the GWAS in populations A and B.

### Adverse events measurement

We provided internet-based questionnaires about the COVID-19 vaccine to the study subjects. First, they were asked about the manufacturer name of the COVID-19 vaccine they were shot. Next, they answered the local or systemic reactions they had after the first and/or second COVID-19 vaccination. The questionnaire provided 51 options for adverse events that are reported to occur with a frequency of 0.1% or more in Japanese subjects^18^. Detailed information about the questionnaires is provided in Table S1.

### Genome-wide association and meta-analysis

The association between genotype dosage and the occurrence of COVID-19 vaccine adverse events was examined using a logistic regression model under the assumption of additive genetic effects. For each population, GWAS was performed with adjustment for age and sex using PLINK (version 2.00a3).

We combined the statistical data from both populations using a fixed-effects model and the inverse-variance weighting method with METAL software^19^ (version 2011-03-25). Variants achieving genome-wide significance (*p* < 5.0 × 10^−8^) in the meta-analysis were considered to be associated with the occurrence of COVID-19 vaccine adverse events.

## Results and Discussion

### Study subjects and occurrence of COVID-19 vaccine adverse events

The data was analyzed independently in the two groups established according to genotyping chips (population A and B), because the analyzed SNP sets were very different between the chips. The characteristics of the subjects are presented in Table 1. The population vaccinated with mRNA-1273 vaccine was older and had a greater prevalence of females, when compared to the population vaccinated with BNT162b1. This difference may be a result of the Japanese vaccination circumstance where the BNT162b1 vaccine was approved at first and administered to vaccination priority targets, such as elderly people and health care workers. The occurrence of each adverse event following the COVID-19 vaccine is shown in Table S2. The occurrence of systemic reactions was more prevalent after the 2^nd^ vaccination dose than after the 1^st^ vaccination (44% and 71% in BNT162b1, 57% and 96% in mRNA-1273, for 1^st^, and 2^nd^ doses, respectively). This is consistent with previous reports^8,20^. Compared to the report from the Japanese Ministry of Health, Labour, and Welfare^18^, most of the adverse events had occurrences with a range of difference of < 5%; 88% and 82% for BNT162b1 vaccine, and 79% and 62% for mRNA-1273 vaccine, for 1^st^ and 2^nd^ dose, respectively (Table S2).

**Table 1.** Characteristics of the subjects included in the present study

|  | population A |  |  |  | population B |  |  |  |
| --- | --- | --- | --- | --- | --- | --- | --- | --- |
|  | BNT162b1 vaccine |  | mRNA-1273 vaccine |  | BNT162b1 vaccine |  | mRNA-1273 vaccine |  |
|  | 1<sup>st</sup> dose | 2<sup>nd</sup> dose | 1<sup>st</sup> dose | 2<sup>nd</sup> dose | 1<sup>st</sup> dose | 2<sup>nd</sup> dose | 1<sup>st</sup> dose | 2<sup>nd</sup> dose |
| N | 2395 | 2017 | 1184 | 1028 | 629 | 537 | 337 | 300 |
| female (%) | 47.06 | 47.00 | 40.96 | 41.53 | 43.40 | 42.08 | 37.98 | 36.67 |
| Age (mean ± SD) | 52.43 ± 11.48 | 53.70 ± 11.37 | 45.61 ± 11.26 | 46.08 ± 11.34 | 52.04 ± 11.77 | 53.01 ± 11.94 | 47.36 ± 10.53 | 47.96 ± 10.65 |

SD, standard deviation

### Incidence of COVID-19 vaccine adverse events with respect to sex and age differences

Previous studies^7,20^ reported that women and younger people have a higher risk of experiencing adverse events after COVID-19 vaccination. Consistent with these studies, the present study also showed a higher risk for females (*p*-value < 0.05; 51% and 63% in BNT162b1, 39% and 53% in mRNA-1273, for 1^st^, and 2^nd^ doses, respectively) and younger people (*p*-value < 0.05; 51% and 63% in BNT162b1, 31% and 41% in mRNA-1273, for 1^st^, and 2^nd^ doses, respectively) (Table S3, S4).

### GWAS for COVID-19 vaccine adverse events

We performed GWAS for each population and performed a meta-analysis of adverse events. We identified 14 loci associated with adverse events in response to COVID-19 vaccine at the genome-wide significance level (*p*-value < 5 × 10^−8^), for1^st^ or 2^nd^ dose of BNT162b1 or mRNA-1273 vaccine (Table 2, Table S5). The associations between rs9266082 and higher fever, and rs13279405 and chest pain were found with *p*-value < 0.05, for both BNT162b1 and mRNA-1273 vaccines. However, the other SNPs were differentially associated with BNT162b1 and mRNA-1273 vaccines. Two hypothesis could explain this fact: the genetic susceptibility to adverse events may differ between the two COVID-19 mRNA vaccines; or the populations may have differential statistical power because of differential sample size and adverse event occurrence. In these associated loci, 6p21 (rs551634406, rs183300, rs9266082, rs375726766, rs3135408) was associated with 37.5 °C or higher fever, 38 °C or higher fever, and muscle pain.

**Table 2.** Loci identified by the meta-analysis of COVID-19 vaccine adverse events

| reaction | SNP | CHR | Position | EA | NEA | EAF | dose | BNT162b1 vaccine |  |  | mRNA-1273 vaccine |  |  |
| --- | --- | --- | --- | --- | --- | --- | --- | --- | --- | --- | --- | --- | --- |
|  |  |  |  |  |  |  |  | OR | SE | P | OR | SE | P |
| itching of vaccination site | rs10744866 | 12 | 110104688 | A | T | 0.37 | 1<sup>st</sup> | 0.89 | 0.12 | 0.317 | 0.55 | 0.11 | $2.42 \times 10^{-8}$ |
|  |  |  |  |  |  |  | 2<sup>nd</sup> | 1.10 | 0.11 | 0.404 | 0.88 | 0.11 | 0.239 |
| movement disorder at the vaccination site | rs146922515 | 10 | 23658821 | CA | C | 0.43 | 1<sup>st</sup> | 1.13 | 0.07 | 0.0783 | 1.23 | 0.09 | **0.0286** |
|  |  |  |  |  |  |  | 2<sup>nd</sup> | 1.03 | 0.08 | 0.738 | 1.80 | 0.11 | **$3.69 \times 10^{-8}$** |
| internal bleeding in vaccination site | rs1217097 | 8 | 64623330 | A | G | 0.24 | 1<sup>st</sup> | 4.12 | 0.24 | **$1.79 \times 10^{-9}$** | 1.11 | 0.41 | 0.797 |
|  |  |  |  |  |  |  | 2<sup>nd</sup> | 1.85 | 0.34 | 0.0699 | 0.25 | 0.78 | 0.0768 |
| 37.5°C or higher fever | rs551634406 | 6 | 30965800 | (A)<sub>15</sub>TAT | A | 0.49 | 1<sup>st</sup> | 0.90 | 0.11 | 0.354 | 0.96 | 0.11 | 0.703 |
|  |  |  |  |  |  |  | 2<sup>nd</sup> | 0.66 | 0.07 | **$1.09 \times 10^{-9}$** | 0.84 | 0.12 | 0.127 |
| 37.5°C or higher fever | rs183300 | 6 | 33526951 | C | T | 0.46 | 1<sup>st</sup> | 0.97 | 0.11 | 0.741 | 0.91 | 0.10 | 0.359 |
|  |  |  |  |  |  |  | 2<sup>nd</sup> | 0.66 | 0.06 | **$3.39 \times 10^{-11}$** | 0.82 | 0.11 | 0.0736 |
| 38°C or higher fever | rs9266082 | 6 | 31320022 | C | T | 0.39 | 1<sup>st</sup> | 1.31 | 0.21 | 0.201 | 1.39 | 0.16 | **0.0448** |
|  |  |  |  |  |  |  | 2<sup>nd</sup> | 1.60 | 0.08 | **$4.59 \times 10^{-9}$** | 1.13 | 0.09 | 0.144 |
| 38°C or higher fever | rs375726766 | 6 | 33335716 | C | CA | 0.36 | 1<sup>st</sup> | 1.04 | 0.23 | 0.856 | 0.90 | 0.18 | 0.556 |
|  |  |  |  |  |  |  | 2<sup>nd</sup> | 0.60 | 0.09 | **$3.67 \times 10^{-8}$** | 0.98 | 0.09 | 0.855 |
| peripheral coolness | rs10205263 | 2 | 221076324 | C | T | 0.090 | 1<sup>st</sup> | 13.09 | 0.44 | **$7.30 \times 10^{-9}$** | 1.31 | 0.93 | 0.771 |
|  |  |  |  |  |  |  | 2<sup>nd</sup> | 3.61 | 0.41 | **0.00180** | 0.37 | 0.56 | 0.0777 |
| dizzy | rs67053119 | 8 | 51750913 | T | A | 0.075 | 1<sup>st</sup> | 0.52 | 0.63 | 0.300 | 1.25 | 0.63 | 0.726 |
|  |  |  |  |  |  |  | 2<sup>nd</sup> | 0.93 | 0.38 | 0.851 | 5.84 | 0.32 | **$2.94 \times 10^{-8}$** |
| chest pain | rs13279405 | 8 | 12986585 | T | C | 0.056 | 1<sup>st</sup> | 17.29 | 0.51 | **$2.69 \times 10^{-8}$** | 7.87 | 0.68 | **0.00225** |
|  |  |  |  |  |  |  | 2<sup>nd</sup> | 5.66 | 0.56 | **0.00210** | 4.38 | 1.02 | 0.146 |
| abdominal discomfort | rs57177321 | 11 | 33988111 | T | C | 0.10 | 1<sup>st</sup> | 7.63 | 0.36 | **$2.30 \times 10^{-8}$** | 0.71 | 0.91 | 0.705 |
|  |  |  |  |  |  |  | 2<sup>nd</sup> | 1.94 | 0.50 | 0.182 | 0.43 | 0.82 | 0.301 |
| joint pain | rs34086990 | 8 | 81791475 | A | AT | 0.19 | 1<sup>st</sup> | 0.72 | 0.31 | 0.284 | 3.78 | 0.24 | **$3.94 \times 10^{-8}$** |
|  |  |  |  |  |  |  | 2<sup>nd</sup> | 1.07 | 0.18 | 0.708 | 1.34 | 0.18 | 0.0962 |
| muscle pain | rs3135408 | 6 | 33275013 | C | T | 0.36 | 1<sup>st</sup> | 0.82 | 0.09 | **0.0392** | 0.84 | 0.11 | 0.111 |
|  |  |  |  |  |  |  | 2<sup>nd</sup> | 0.56 | 0.10 | **$1.13 \times 10^{-8}$** | 0.88 | 0.10 | 0.222 |
| urticaria | rs2274569 | 1 | 100435079 | C | T | 0.064 | 1<sup>st</sup> | 3.82 | 0.51 | **0.00811** | 2.94 | 0.79 | 0.170 |
|  |  |  |  |  |  |  | 2<sup>nd</sup> | 11.78 | 0.45 | **$3.15 \times 10^{-8}$** | 1.91 | 1.23 | 0.600 |

Loci that reached genome-wide significance after meta-analysis at the 1<sup>st</sup> or 2<sup>nd</sup> dose of BNT162b1 or mRNA-1273 vaccine. CHR, chromosome; EA, effect allele; NEA, non-effect allele; EAF, effect allele frequency; OR, odds ratio of effect allele; SE, standard error for beta of effect allele; P, *p*-value

The annotations of the associated SNPs are shown in Tables 3^21,22^. The occurrence of adverse events in response to COVID-19 vaccines was previously reported to differ among ethnicities. The prevalence of fatigue as a reaction to BNT162b2 vaccine was 59% and 69%, and fever was 16% and 38%, in European and Japanese populations, respectively^8,18,20^. The prevalence of fatigue as a reaction to mRNA-1273 vaccine was 68% and 80%, and fever was 17% and 77%, in European and Japanese populations, respectively^8,18,20^. In our study, the allele frequencies for eight out of the 14 SNPs (rs10744866, rs146922515, rs551634406, rs183300, rs375726766, rs13279405, rs34086990, rs3135408) differed by more than 10% between European and Japanese populations, suggesting the possibility that the difference in genetic backgrounds may influence the occurrence of adverse events in response to COVID-19 vaccines. These loci, especially 6p21, were associated with the expression of many genes according to GTEx^22^. These genes were suggested to influence the mechanism of action of the COVID-19 vaccine. *HLA* genes’ *(HLA-B, C, DPA1*) mRNA expression differed among the genotypes of the associated loci. *HLA* alleles were also reported to be associated with adverse events following COVID-19 mRNA vaccination^10^. In fact, *HLA* genes have been reported to be associated with the occurrence of adverse events after administration of various vaccines^23^ and drugs^24^. Regarding other genes, increased *NOTCH4* expression in circulating regulatory T cells in COVID-19 patients was associated with disease severity and predicted mortality^25^. The expression of *RPS18* was previously found to be increased in isolated T-cells on stimulation with the live influenza virus^26^. A SNP in the *BAK1* and A haplotype of *MICB* was associated with dengue hemorrhagic fever caused by dengue virus^27,28^. A SNP in *PSORS1C1* was associated with allopurinol-induced severe adverse reactions^29,30^. SNPs in *HSP70, TAPBP*, and *WDR46* were found to be associated with aspirin-exacerbated respiratory disease^31–33^.

**Table 3.** Allele frequency and expression quantitative trait locus (eQTL) genes for loci associated with COVID-19 vaccine adverse events

| SNP | associated reaction | location | allele frequency |  | eQTL genes |
| --- | --- | --- | --- | --- | --- |
|  |  |  | European | Japanese |  |
| rs10744866 | itching of vaccination site | 12q24 | 0.20 | 0.37 | *MVK, FOXN4, KCTD10* |
| rs146922515 | movement disorder at the vaccination site | 10p12 | 0.35 | 0.43 | None in GTEx |
| rs1217097 | internal bleeding in vaccination site | 8q12 | 0.28 | 0.24 | *RP11-579E24.2, LINC01289* |
| rs551634406 | 37.5°C or higher fever | 6p21 | 1.00 | 0.49 | None in GTEx |
| rs183300 | 37.5°C or higher fever | 6p21 | 0.65 | 0.46 | *BAK1, COL11A2, DAXX, IP6K3* |
| rs9266082 | 38°C or higher fever | 6p21 | 0.30 | 0.39 | *C4B, CCHCR1, CSNK2B, HCG22, HCG27, HLA-B, HLA-C, HSPA1B, MICB, MIR6891, NOTCH4, POU5F1, PSORS1C1, PSORS1C2, RNF5, SFTA2, TCF19, USP8P1, VARS2, VWA7, WASF5P, XXbac-BPG181B23.7, XXbac-BPG248L24.12, XXbac-BPG299F13.17* |
| rs375726766 | 38°C or higher fever | 6p21 | 0.60 | 0.36 | None in GTEx |
| rs10205263 | peripheral coolness | 2q35 | 0.071 | 0.09 | No eQTL genes |
| rs67053119 | dizzy | 8q11 | 0.12 | 0.075 | No eQTL genes |
| rs13279405 | chest pain | 8p22 | 0.24 | 0.056 | No eQTL genes |
| rs57177321 | abdominal discomfort | 11p13 | 0.072 | 0.1 | No eQTL genes |
| rs34086990 | joint pain | 8q21 | 0.042 | 0.19 | None in GTEx |
| rs3135408 | muscle pain | 6p21 | 0.54 | 0.36 | *B3GALT4, BAK1, DAXX, HCG24, HCG25, HLA-DPA1, RGL2, RPS18, TAPBP, WDR46, ZBTB22* |
| rs2274569 | urticaria | 1p21 | 0.076 | 0.064 | *DBT, LRRC39, MFSD14A, RTCA, SASS6, SLC35A3, TRMT13* |

European allele frequency, allele frequency of European (non-Finnish) in gnomAD v2,1,1; Japanese allele frequency, allele frequency in this study; eQTL genes, SNPs associated with COVID-19 vaccine adverse events were associated with gene expression in GTEx ( $p$ -value <0.0005).

### Limitations

This study has some limitations. Initially, we included 51 types of adverse events, but multiple test corrections were not performed. Therefore, the associated loci that we identified can include false positives, and thus, replicate studies are required. Second, our data on the adverse events was based on web-based self-reports and might have been affected by recall bias. However, the occurrence of adverse events was similar to the large-scale survey performed before in Japan^18^. Thus, our adverse event data is thought to have some reliability. Finally, our GWAS was based only on the Japanese population. Therefore, our results may not be directly applicable to other ethnicities.

## Conclusions

In this study, we performed GWAS for adverse events following COVID-19 vaccination. To the extent of our knowledge, this work represents the first of its kind focusing on East Asian populations. We identified 14 loci associated with adverse effects of COVID-19 vaccines, in the Japanese population. We discovered that the genetic background was in fact associated with susceptibility to adverse events following COVID-19 vaccination. Our results may enable one to prepare for and manage adverse events on the basis of their susceptibility to the occurrence of adverse events. Furthermore, we obtained valuable basic data that can be used for investigating the mechanism of action of COVID-19 vaccines.

## Data Availability

All data analyzed during this study are included in this published article and its additional files. Other data are available from the authors upon reasonable request.

## Acknowledgements

We are grateful to the “Genequest ALL” and “Euglena MyHealth” participants included in this study. We thank Euglena Co., Ltd. for making this study possible.

## Author contributions

K.S. and S.T. designed the experiment. S.N., K.K., M.C., H.K., and K.T. created the questionnaire about the COVID-19 vaccine and reviewed the data. S.N. performed the statistical analyses. S.N. and K.S. wrote the manuscript. H.K. and K.T. contributed to the interpretation of the results and critically reviewed the manuscript. All authors commented on and approved the manuscript.

## Funding

This work was supported by internal funding from Genequest, Inc.

## Competing interests

S.N., K.K, and M.C. are employees of Genequest Inc.; K.S. and S.T. are board members of Genequest Inc.; H.K. and K.T. declare no competing interests.

## Notes

### Author Declarations

This study was approved by the Ethics Committee of Genequest Inc. (IRB no. 2021-0633-4) and Tohoku University Graduate School of Medicine (IRB no. 2021-1-469).

## References

1. Hu B, Guo H, Zhou P, Shi ZL. Characteristics of SARS-CoV-2 and COVID-19. Nat Rev Microbiol 2021;19(3):141–154. doi:10.1038/s41579-020-00459-7

2. COVID-19 vaccine tracker and landscape. World Health Organization (WHO). Accessed November 5, 2021. Posted online https://www.who.int/publications/m/item/draft-landscape-of-covid-19-candidate-vaccines

3. Novel coronavirus vaccines. Official Website of the prime minister of Japan and His cabinet. Accessed November 29, 2021. Posted online https://www.kantei.go.jp/jp/headline/kansensho/vaccine.html

4. Chapin-Bardales J, Gee J, Myers T. Reactogenicity following receipt of mRNA-based COVID-19 vaccines. JAMA 2021;325(21):2201–2202. doi:10.1001/jama.2021.5374

5. Shimabukuro T, Nair N. Allergic reactions including anaphylaxis after receipt of the first dose of Pfizer-BioNTech COVID-19 vaccine. JAMA 2021;325(8):780–781. doi:10.1001/jama.2021.0600

6. Klimek L, Bergmann KC, Brehler R, et al. Practical handling of allergic reactions to COVID-19 vaccines: A position paper from German and Austrian Allergy Societies AeDA, DGAKI, GPA and ÖGAI. Allergo J Int 2021;30(3):1–17. doi:10.1007/s40629-021-00165-7

7. Menni C, Klaser K, May A, et al. Vaccine side-effects and SARS-CoV-2 infection after vaccination in users of the COVID Symptom Study app in the UK: A prospective observational study. Lancet Infect Dis 2021;21(7):939–949. doi:10.1016/S1473-3099(21)00224-3

8. Polack FP, Thomas SJ, Kitchin N, et al. Safety and efficacy of the BNT162b2 mRNA Covid-19 vaccine. N Engl J Med 2020;383(27):2603–2615. doi:10.1056/NEJMoa2034577

9. Gargano JW, Wallace M, Hadler SC, et al. Use of mRNA COVID-19 vaccine after reports of myocarditis among vaccine recipients: Update from the Advisory Committee on Immunization Practices — United States, June 2021. MMWR Morb Mortal Wkly Rep 2021;70(27):977–982. doi:10.15585/mmwr.mm7027e2

10. The genetics behind the different reactions to COVID-19 vaccines. 23andMe blog. Published 9 November, 2021. Accessed November 24, 2021. Posted online https://blog.23andme.com/23andme-research/reaction-to-covid-vaccine/

11. Bolze A, Neveux I, Barrett KMS, et al. HLA-A*03:01 Is Associated with Increased Risk of Fever, Chills, and More Severe Reaction to Pfizer-BioNTech COVID-19 Vaccination. 2021:2021.11.16.21266408. doi:10.1101/2021.11.16.21266408

12. Second-generation PLINK: Rising to the challenge of larger and richer datasets | GigaScience | Oxford. Academic. Accessed November 17, 2021. Posted online https://academic.oup.com/gigascience/article/4/1/s13742-015-0047-8/2707533

13. Purcell S, Neale B, Todd-Brown K, Thomas L, Ferreira MA, Bender D, Maller J, Sklar P, de Bakker PI, Daly MJ, Sham PC. PLINK: A tool set for whole-genome association and population-based linkage analyses. Am J Hum Genet 2007;81(3):559–575

14. Price AL, Patterson NJ, Plenge RM, Weinblatt ME, Shadick NA, Reich D. Principal components analysis corrects for stratification in genome-wide association studies. Nat Genet 2006;38(8):904–909. doi:10.1038/ng1847

15. Loh PR, Danecek P, Palamara PF, et al. Reference-based phasing using the Haplotype Reference Consortium panel. Nat Genet 2016;48(11):1443–1448. doi:10.1038/ng.3679

16. Das S, Forer L, Schönherr S, et al. Next-generation genotype imputation service and methods. Nat Genet 2016;48(10):1284–1287. doi:10.1038/ng.3656

17. 1000 Genomes Project Consortium, Auton A, Brooks LD, Durbin RM, Garrison EP, Kang HM, Korbel JO, Marchini JL, McCarthy S, McVean GA, Abecasis GR. A global reference for human genetic variation. Nature 2015;526(7571):68–74. doi:10.1038/nature15393

18. Health status survey after COVID-19 vaccination. Ministry of Health, Labour and Welfare Web site. Accessed November 12, 2021. Posted online https://www.mhlw.go.jp/stf/seisakunitsuite/bunya/vaccine_kenkoujoukyoutyousa.html

19. Willer CJ, Li Y, Abecasis GR. METAL: Fast and efficient meta-analysis of genomewide association scans. Bioinformatics 2010;26(17):2190–2191. doi:10.1093/bioinformatics/btq340

20. Baden LR, El Sahly HME, Essink B, et al. Efficacy and safety of the mRNA-1273 SARS-CoV-2 vaccine. N Engl J Med 2021;384(5):403–416 doi:10.1056/NEJMoa2035389

21. Karczewski KJ, Francioli LC, Tiao G, et al. The mutational constraint spectrum quantified from variation in 141,456 humans. Nature 2020;581(7809):434–443. doi:10.1038/s41586-020-2308-7

22. GTEx Consortium. Human genomics. The Genotype-Tissue Expression (GTEx) pilot analysis: Multitissue gene regulation in humans. Science 2015;348(6235):648–660. doi:10.1126/science.1262110

23. Borba V, Malkova A, Basantsova N, et al. Classical examples of the concept of the ASIA syndrome. Biomolecules 2020;10(10):1436. doi:10.3390/biom10101436

24. Fan WL, Shiao MS, Hui RC, Su SC, Wang CW, Chang YC, Chung WH, et al. HLA association with drug-induced adverse reactions. J Immunol Res 2017;2017:3186328. doi:10.1155/2017/3186328

25. Harb H, Benamar M, Lai PS, et al. Notch4 signaling limits regulatory T-cell-mediated tissue repair and promotes severe lung inflammation in viral infections. Immunity 2021;54(6):1186–1199.e7. doi:10.1016/j.immuni.2021.04.002

26. Roy JG, McElhaney JE, Verschoor CP. Reliable reference genes for the quantification of mRNA in human T-cells and PBMCs stimulated with live influenza virus. BMC Immunol 2020;21(1):4. doi:10.1186/s12865-020-0334-8

27. Dang TN, Naka I, Sa-Ngasang A, et al. Association of BAK1 single nucleotide polymorphism with a risk for dengue hemorrhagic fever. BMC Med Genet 2016;17(1):43. doi:10.1186/s12881-016-0305-3

28. Luangtrakool P, Vejbaesya S, Luangtrakool K, et al. Major histocompatibility complex Class I chain-related A and B (MICA and MICB) gene, allele, and haplotype associations with dengue infections in ethnic Thais. J Infect Dis 2020;222(5):840–846. doi:10.1093/infdis/jiaa134

29. Cheng L, Sun B, Xiong Y, et al. The minor alleles HCP5 rs3099844 A and PSORS1C1 rs3131003 G are associated with allopurinol-induced severe cutaneous adverse reactions in Han Chinese: A multicentre retrospective case-control clinical study. Br J Dermatol 2018;178(3):e191–e193. doi:10.1111/bjd.16151

30. Maekawa K, Nishikawa J, Kaniwa N, et al. Development of a rapid and inexpensive assay for detecting a surrogate genetic polymorphism of HLA-B*58:01: A partially predictive but useful biomarker for allopurinol-related Stevens-Johnson syndrome/toxic epidermal necrolysis in Japanese. Drug Metab Pharmacokinet 2012;27(4):447–450. doi:10.2133/dmpk.dmpk-11-nt-120

31. Kooti W, Abdi M, Malik YS, et al. Association of CYP2C19 and HSP70 genes polymorphism with aspirin-exacerbated respiratory disease in a Kurd population. Endocr Metab Immune Disord Drug Targets 2020;20(2):256–262. doi:10.2174/1872214812666190527104329

32. Cho SH, Park JS, Park BL, Bae DJ, Uh ST, Kim MK, Choi IS, Shin HD, Park CSAssociation analysis of tapasin polymorphisms with aspirin-exacerbated respiratory disease in asthmatics. Pharmacogenet Genomics 2013;23(7):341–348. doi:10.1097/FPC.0b013e328361d4bb

33. Pasaje CFA, Bae JS, Park BL, et al. WDR46 is a genetic risk factor for aspirin-exacerbated respiratory disease in a Korean population. Allergy Asthma Immunol Res 2012;4(4):199–205. doi:10.4168/aair.2012.4.4.199

